# Impact of Cardiovascular-Kidney-Metabolic Burden on Echocardiographic Remodeling and Cardiovascular Outcomes in Hypertension

**DOI:** 10.64898/2026.10.21.26366110

**Authors:** Hyue Mee Kim, Minjung Bak, Jiesuck Park, Hong-Mi Choi, Yeonyee E. Yoon, Goo-Yeong Cho, In-Chang Hwang

## Abstract

**Aims:** Cardiovascular-kidney-metabolic (CKM) syndrome highlights the interrelationship between metabolic, renal, and cardiovascular disorders. However, whether CKM burden influences longitudinal cardiac structural and functional recovery and subsequent cardiovascular outcomes remains unclear.

**Methods:** We studied 1,600 patients from the STRATS-HHD registry who underwent echocardiography at baseline and 6–18 months after treatment initiation. CKM burden was defined by obesity, diabetes, chronic kidney disease, and severe hypertension (0–1, 2, or ≥3 components). The primary outcome was cardiovascular death or heart failure hospitalization (HHF).

**Results:** CKM burden was 0–1 in 973 patients (60.8%), 2 in 440 (27.5%), and ≥3 in 187 (11.7%). Higher burden was associated with worse baseline cardiac abnormalities and increased primary outcome risk (adjusted HR, 3.09; 95% CI, 1.92–4.98 for ≥3 vs 0–1; adjusted HR, 1.53; 95% CI, 1.27–1.84 per +1 CKM component). The prognostic association persisted after additional adjustment for hepatic steatosis index-defined metabolic dysfunction-associated steatotic liver disease. Despite overall improvement in most echocardiographic parameters after treatment, patients with CKM burden ≥3 showed less favorable E/e′, LV global longitudinal strain (GLS), and LA stiffness index at follow-up. Normalization of these parameters at follow-up was associated with substantially lower subsequent cardiovascular risk. Mediation analyses suggested that echocardiographic abnormalities partially mediated the association between high CKM burden and the primary outcome.

**Conclusion:** CKM burden identifies hypertensive patients with less favorable cardiac function after treatment and increased cardiovascular risk. These findings highlight the association of CKM burden with cardiac functional recovery and prognosis in patients with hypertension.

**Graphical Abstract:** In patients with hypertension, higher CKM burden was associated with more pronounced cardiac abnormalities, less favorable cardiac functional status after antihypertensive treatment, and increased cardiovascular risk. Normalization of functional echocardiographic parameters was associated with lower subsequent cardiovascular risk.

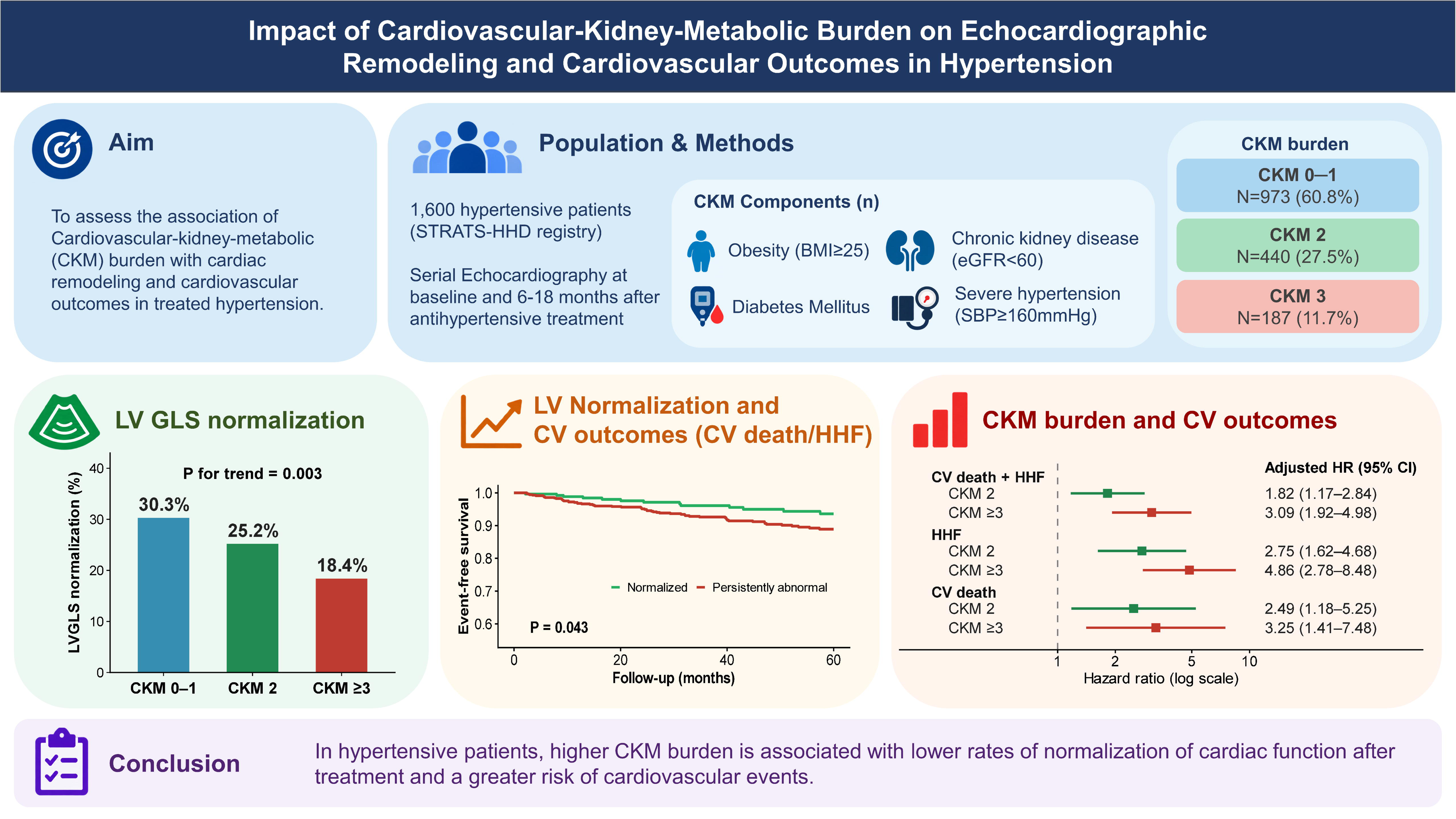

## Introduction

The cardiovascular-kidney-metabolic (CKM) syndrome framework, proposed by the American Heart Association in 2023, conceptualizes metabolic dysfunction, chronic kidney disease, and cardiovascular disease as interconnected processes along a shared continuum rather than as separate entities.^1,2^ More advanced CKM stages have been associated with substantially increased cardiovascular (CV) risk, and recent studies have further shown that higher CKM stages are linked to unfavorable cardiac structural and functional alterations, suggesting that the framework captures cardiac phenotype, as well as future risk, with CV manifestations emerging well before overt clinical disease.^3–6^

However, CKM syndrome reflects the coexistence of multiple cardiometabolic abnormalities rather than a single disease process,^1^ and individuals within the same stage may carry markedly different burdens of obesity, hypertension, diabetes, and kidney dysfunction.^4,7,8^ Thus, the cumulative burden of CKM components may capture heterogeneity within stages and provide complementary information on cardiac phenotype, remodeling, and risk beyond categorical staging.

Hypertension is one of several metabolic risk factors within the CKM framework. Chronic pressure overload can lead to cardiac structural and functional remodeling, which may be partially reversible with antihypertensive treatment and blood-pressure reduction.^9,10^ However, whether higher CKM burden is associated with less favorable cardiac recovery following antihypertensive treatment, and whether such recovery is associated with subsequent CV risk, remains largely unknown in hypertensive patients. Therefore, we investigated the association of CKM burden with baseline echocardiographic phenotype, longitudinal echocardiographic remodeling, and CV outcomes in a large cohort of patients with hypertension. We further examined whether recovery of echocardiographic abnormalities differed according to CKM burden and whether such recovery was associated with subsequent CV outcomes.

## Methods

### Study population

This study was conducted using data from the Strain for Risk Assessment and Therapeutic Strategies in Patients With Hypertensive Heart Disease (STRATS-HHD) registry, a multicenter observational cohort of patients with essential hypertension.^9–13^ Consecutive patients who underwent comprehensive transthoracic echocardiography at the time of hypertension diagnosis and a follow-up examination after 6–18 months of antihypertensive treatment were enrolled from Seoul National University Bundang Hospital and Chung-Ang University Hospital between 2006 and 2021. Patients were excluded if they had specific cardiomyopathies (dilated, hypertrophic, restrictive, ischemic, stress-induced, Fabry, or mitochondrial cardiomyopathy), moderate or greater valvular heart disease, end-stage renal disease, previous cardiac surgery, secondary hypertension, or other cardiovascular conditions associated with left ventricular (LV) hypertrophy (LVH). After application of the exclusion criteria, 1,872 patients remained eligible. Among them, 272 were excluded because archived echocardiographic studies were not suitable for longitudinal analysis owing to limitations in image storage and retrieval from earlier study years. The final study population consisted of 1,600 patients (**Supplementary Figure S1**).

The study protocol complied with the Declaration of Helsinki and was approved by the institutional review boards of the participating institutions (Seoul National University Bundang Hospital: IRB No. B-2206-762-102; Chung-Ang University Hospital: IRB No. 2205-014-19419).

The requirement for informed consent was waived owing to the retrospective nature of the study.

### Definition of CKM burden

To quantify the cumulative burden of cardiovascular-kidney-metabolic abnormalities, four CKM components reflecting the obesity, glycemic, renal, and blood pressure (BP) domains of CKM health were identified at baseline, consistent with prior approaches using CKM component counts to capture disease burden beyond categorical staging.^4^ These components were obesity (body mass index ≥25 kg/m² according to Asian-specific criteria^14^), diabetes mellitus (prior diagnosis or treatment), chronic kidney disease (estimated glomerular filtration rate <60 mL/min/1.73 m²), and severe hypertension (baseline systolic BP [SBP] ≥160 mmHg). Each component was assigned one point, yielding a total CKM burden score ranging from 0 to 4. Patients were subsequently categorized into three groups according to CKM burden: 0–1, 2, and ≥3 components.

To examine the potential role of metabolic dysfunction–associated steatotic liver disease (MASLD), which is an integrative indicator of metabolic burden,^15^ hepatic steatosis was estimated using the hepatic steatosis index (HSI), calculated as 8 × (ALT/AST ratio) + body mass index (+2 if female and +2 if diabetes mellitus). An HSI >36 was considered indicative of hepatic steatosis and was used to define HSI-based MASLD.^16^

### Echocardiographic Analysis

LV dimensions, wall thickness, relative wall thickness, LV mass index (LVMI), left atrial (LA) volume index, transmitral Doppler parameters, and tissue Doppler velocities were measured according to contemporary guideline recommendations.^17^ LV global longitudinal strain (LVGLS) and LA reservoir strain (LASr) were quantified using a fully automated artificial intelligence– based software platform (Sonix Health Workstation version 2.0; Ontact Health Inc., Korea).^13,18–20^ LVGLS was calculated as the average peak longitudinal strain from the apical 4-, 2-, and 3-chamber views and expressed as an absolute value. LASr was derived as the peak positive atrial strain during ventricular systole from apical 4- and 2-chamber views. Further technical details and validation have been described previously.^19^ The LA stiffness index was calculated as the ratio of E/e′ to LASr. Longitudinal changes in echocardiographic parameters were assessed by comparing baseline and follow-up measurements. For analyses of echocardiographic normalization, normal values were defined as LVGLS ≥18%, E/e′ <14, and LA stiffness index <0.63. Thresholds for LVGLS and E/e′ were based on established reference criteria,^21,22^ whereas the LA stiffness index cutoff was derived from our previous study.^23^ Normalization was defined as a change from an abnormal value at baseline to a normal value at follow-up.

### Outcomes

The primary outcome was a composite of CV death and hospitalization for heart failure (HHF). Secondary analyses were performed for each individual component of the primary outcome. HHF was defined as an unplanned hospital admission due to worsening heart failure requiring intravenous therapy. CV death was defined as death resulting from myocardial infarction, heart failure, stroke, sudden cardiac death, or other CV causes. For analyses evaluating the prognostic significance of longitudinal echocardiographic changes and recovery, follow-up time was calculated from the date of the follow-up echocardiographic examination, and only events occurring thereafter were included. Clinical outcomes were ascertained through review of electronic medical records, structured telephone follow-up, and linkage with national mortality databases.^9,13^

### Statistical analysis

Continuous variables are expressed as mean ± standard deviation, and categorical variables as frequencies and percentages. Group differences were assessed using Student’s t-test or analysis of variance for continuous variables and the chi-square test for categorical variables, as appropriate. For serial echocardiographic measurements, within-group changes from baseline to follow-up were assessed using the paired t-test. Between-group differences in follow-up values were evaluated using analysis of covariance (ANCOVA), with CKM burden group as the grouping factor and the corresponding baseline value, age, sex, and change in SBP (ΔSBP) as covariates. These covariates were selected to account for baseline echocardiographic status, demographic factors, and the degree of BP reduction during follow-up. Event-free survival was estimated with the Kaplan–Meier method and compared across CKM burden groups using the log-rank test. Cox proportional hazards regression was used to examine associations between CKM burden group and clinical outcomes. To assess whether the association between CKM burden and CV outcomes was predominantly driven by any single component, sensitivity analyses were performed by sequentially excluding each CKM component from the burden score. Mediation analysis was performed using the mediation package in R to evaluate the extent to which echocardiographic parameters (LVGLS, LVMI, E/e’, LASr, and LA stiffness index) mediated the association between CKM burden (≥3 vs 0–2 components) and the primary outcome. The outcome was modeled using logistic regression, with adjustment for age and sex. Average causal mediation effects (ACME), direct effects, and proportions mediated were estimated using nonparametric bootstrapping (5,000 resamples). A two-sided P value <0.05 was considered statistically significant. All analyses were performed using R software (version 4.2.1; R Foundation for Statistical Computing, Vienna, Austria).

## Results

### Baseline Characteristics According to CKM Burden

Among 1,600 patients included in the analysis, 973 (60.8%) were classified as CKM burden 0–1, 440 (27.5%) as CKM burden 2, and 187 (11.7%) as CKM burden ≥3. The prevalence of individual CKM components was 48.7% for obesity, 28.9% for diabetes mellitus, 22.4% for chronic kidney disease, and 31.1% for severe hypertension. Higher CKM burden was associated with more adverse clinical, laboratory, and echocardiographic profiles (**Table 1**).

**Table 1.**
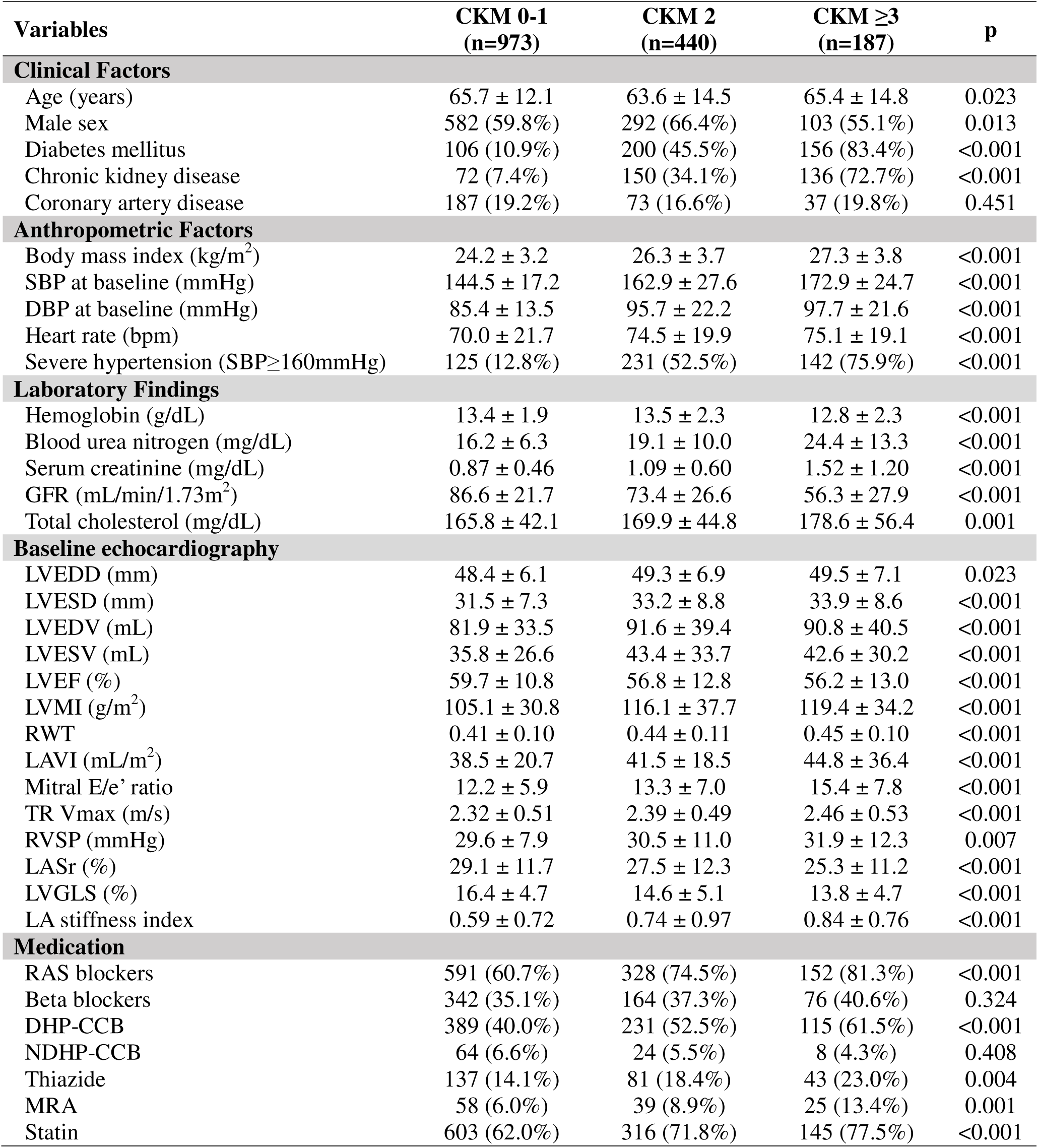
Baseline characteristics.

On baseline echocardiography, patients with higher CKM burden demonstrated progressively greater cardiac structural and functional abnormalities. LV dimensions and volumes increased across CKM burden groups, while LV ejection fraction progressively decreased. Markers of cardiac remodeling, including LVMI and LAVI, were higher in patients with greater CKM burden. Indices of diastolic dysfunction and filling pressure, including E/e′, showed a stepwise increase, accompanied by higher pulmonary artery systolic pressure. Strain-derived parameters also progressively worsened with increasing CKM burden, with lower LVGLS and LASr and higher LA stiffness index across CKM burden groups (**Table 1**).

### Longitudinal Changes in Echocardiographic Parameters According to CKM Burden

Longitudinal changes in echocardiographic parameters across CKM burden groups are illustrated in **Figure 1**. Most echocardiographic parameters showed significant within-group improvement from baseline to follow-up, although not all parameters reached statistical significance across every CKM burden group. Observed follow-up values and ANCOVA-adjusted estimated marginal means, representing predicted follow-up values after accounting for baseline differences, age, sex, and change in systolic blood pressure, are presented in **Supplementary Table S1**. After adjustment, follow-up E/e′ ratio and LA stiffness index remained significantly higher, whereas LVGLS remained significantly lower, in patients with CKM burden ≥3 compared with those with CKM burden 0–1. In contrast, follow-up values of LVMI, LAVI, and LASr were comparable across CKM burden groups.

**Figure 1.**
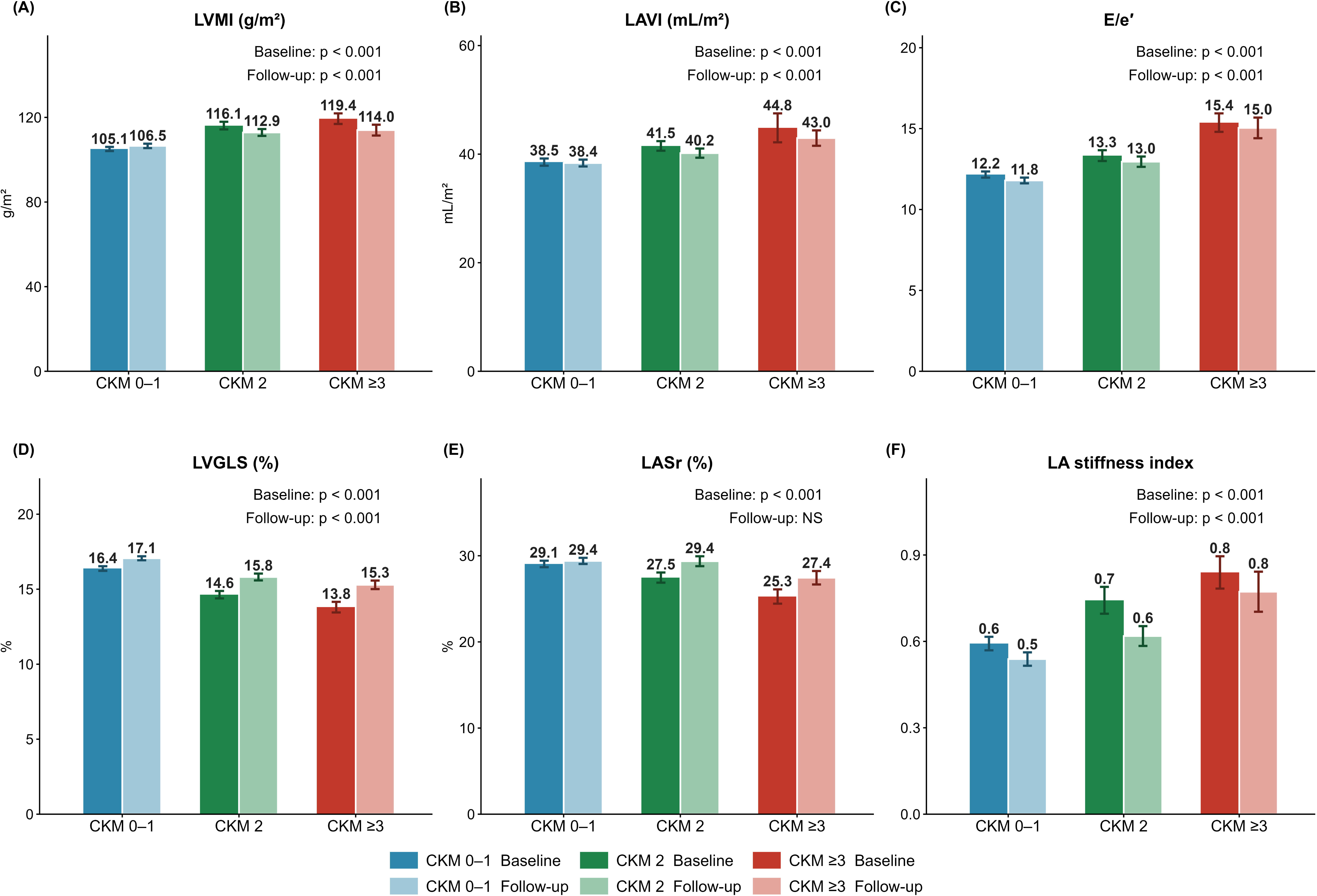
Longitudinal Changes in Echocardiographic Parameters According to CKM Burden Echocardiographic parameters at baseline and follow-up according to CKM burden. (A) Left ventricular mass index (LVMI), (B) left atrial volume index (LAVI), (C) E/e′ ratio, (D) left ventricular global longitudinal strain (LVGLS), (E) left atrial reservoir strain (LASr), and (F) left atrial stiffness index. Alt text: Six-panel figure comparing echocardiographic parameters at baseline and follow-up across three CKM burden groups. Panels show LVMI, LAVI, E/e′ ratio, LVGLS, LASr, and LA stiffness index.

### Prognostic Significance of Echocardiographic Recovery

Given that LVGLS, E/e′ ratio, and LA stiffness index showed less favorable recovery in patients with CKM burden ≥3, we examined whether normalization of these parameters was associated with subsequent CV outcomes (**Table 2**). Kaplan-Meier analysis demonstrated better event-free survival in patients who achieved normalization compared with those with persistently abnormal values across all three parameters (**Figure 2**). Patients who achieved LVGLS normalization had a 47% lower risk of the composite outcome compared with those with persistently impaired LVGLS (adjusted HR, 0.53; 95% CI, 0.30–0.92; p=0.025). Normalization of E/e′ (adjusted HR, 0.20; 95% CI, 0.10–0.40; p<0.001) and LA stiffness index (adjusted HR, 0.11; 95% CI, 0.04– 0.31; p<0.001) were associated with lower CV risk. These trends were maintained across the CKM burden groups (**Table 2**). Notably, the proportion of patients achieving LVGLS normalization progressively decreased with increasing CKM burden (30.3%, 25.2%, and 18.4% in CKM burden 0–1, 2, and ≥3 groups, respectively; p for trend=0.003). Similar numerical trends were observed for normalization of E/e′ and LA stiffness index, although the between-group differences were not statistically significant.

**Figure 2.**
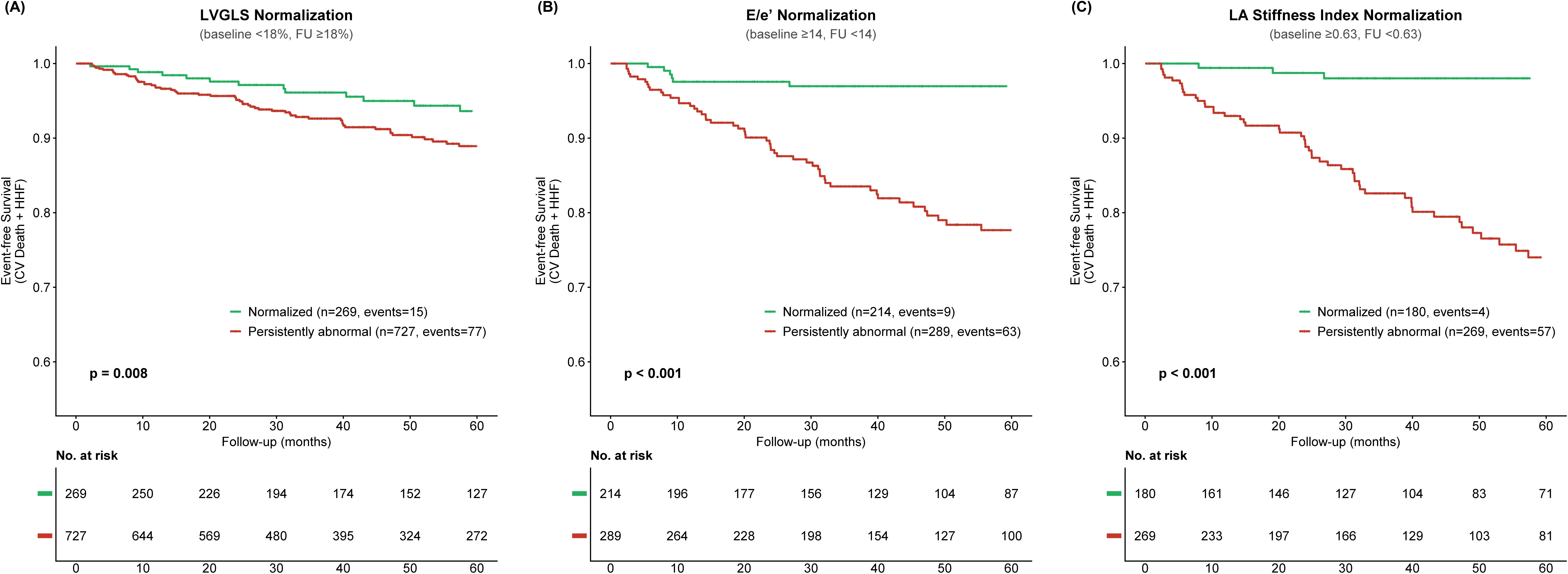
Echocardiographic Normalization and Cardiovascular Outcomes Kaplan–Meier curves for the primary composite outcome according to normalization of echocardiographic parameters among patients with abnormal values at baseline. (A) LVGLS normalization, (B) E/e′ normalization, and (C) LA stiffness index normalization. Alt text: Three Kaplan–Meier curves comparing cardiovascular event-free survival between patients with normalized and persistently abnormal echocardiographic parameters at follow-up. Panels show LVGLS, E/e′ ratio, and LA stiffness index, with higher event-free survival in patients achieving normalization.

**Table 2.** Follow-up echocardiographic normalization among patients with baseline abnormal findings and cardiovascular outcomes.

| CKM Group | Patients achieving normalization | Events in normalized (%) | Events in not normalized (%) | Adjusted HR (95% CI) | P value |
| --- | --- | --- | --- | --- | --- |
| LV GLS normalization (baseline <18% → follow-up ≥18%) |  |  |  |  |  |
| Total | 269 / 996 (27.0%) | 15 (5.6%) | 77 (10.6%) | 0.53 (0.30—0.92) | 0.025 |
| 0–1 | 165 / 545 (30.3%) | 5 (3.0%) | 24 (6.3%) |  |  |
| 2 | 78 / 310 (25.2%) | 5 (6.4%) | 27 (11.6%) |  |  |
| ≥3 | 26 / 141 (18.4%) | 5 (19.2%) | 26 (22.6%) |  |  |
| E/e' normalization (baseline ≥14 → follow-up <14) |  |  |  |  |  |
| Total | 214 / 503 (42.5%) | 9 (4.2%) | 63 (21.8%) | 0.20 (0.10—0.40) | <0.001 |
| 0–1 | 122 / 269 (45.4%) | 2 (1.6%) | 24 (16.3%) |  |  |
| 2 | 61/ 147 (41.5%) | 3 (4.9%) | 20 (23.3%) |  |  |
| ≥3 | 31 / 87 (35.6%) | 4 (12.9%) | 19 (33.9%) |  |  |
| LA stiffness index normalization (baseline ≥0.63 → follow-up <0.63) |  |  |  |  |  |
| Total | 180 / 449 (40.1%) | 4 (2.2%) | 57 (21.2%) | 0.11 (0.04—0.31) | <0.001 |
| 0–1 | 93 / 235 (39.6%) | 2 (2.2%) | 18 (12.7%) |  |  |
| 2 | 59 / 139 (42.4%) | 0 (0.0%) | 23 (28.7%) |  |  |
| ≥3 | 28 / 75 (37.3%) | 2 (7.1%) | 16 (34.0%) |  |  |

Abbreviations: CKM, cardiovascular-kidney-metabolic; E/e', mitral E/e' ratio; LVGLS, left ventricular global longitudinal strain; LASr, left atrial reservoir strain; LA, left atrial.
Normalization defined as: LVGLS, baseline <18% → follow-up ≥18%; E/e', baseline ≥14 → follow-up <14; LA stiffness index, baseline ≥0.63 → follow-up <0.63. LA stiffness index cutoff derived from prior analysis in the same cohort. Adjusted HR: Cox proportional hazards model adjusted for age and sex.

### Association of CKM Burden with Cardiovascular Outcomes

Kaplan-Meier analysis demonstrated a stepwise increase in the incidence of the primary outcome across CKM burden groups. Similar graded associations were observed for HHF and CV death analyzed separately (**Figure 3**). In Cox proportional hazards analyses adjusted for age and sex, increasing CKM burden was independently associated with a higher risk of the primary outcome, HHF, and CV death (**Table 3, Supplementary Figure S2**). These associations remained significant after further adjustment for prior coronary artery disease and stroke. Compared with the CKM burden 0–1 group, CKM burden ≥3 was associated with higher risks of the primary composite outcome (adjusted HR, 3.09; 95% CI, 1.92–4.98; p<0.001), HHF (adjusted HR, 4.86; 95% CI, 2.78–8.48; p<0.001), and CV death (adjusted HR, 3.25; 95% CI, 1.41–7.48; p=0.006). When modeled as a continuous variable, each additional CKM component was associated with a higher risk of the primary composite outcome (adjusted HR, 1.54; 95% CI, 1.28–1.85; p<0.001), HHF (adjusted HR, 1.84; 95% CI, 1.49–2.27; p<0.001), and CV death (adjusted HR, 1.52; 95% CI, 1.10–2.06; p=0.008) (**Table 3, Supplementary Figure S2**).The association between increasing CKM burden and the primary outcome remained robust after sequential exclusion of each individual CKM component (HR per 1-component increase, 1.49–1.67; all p<0.001; **Supplementary Table S2**).

**Figure 3.**
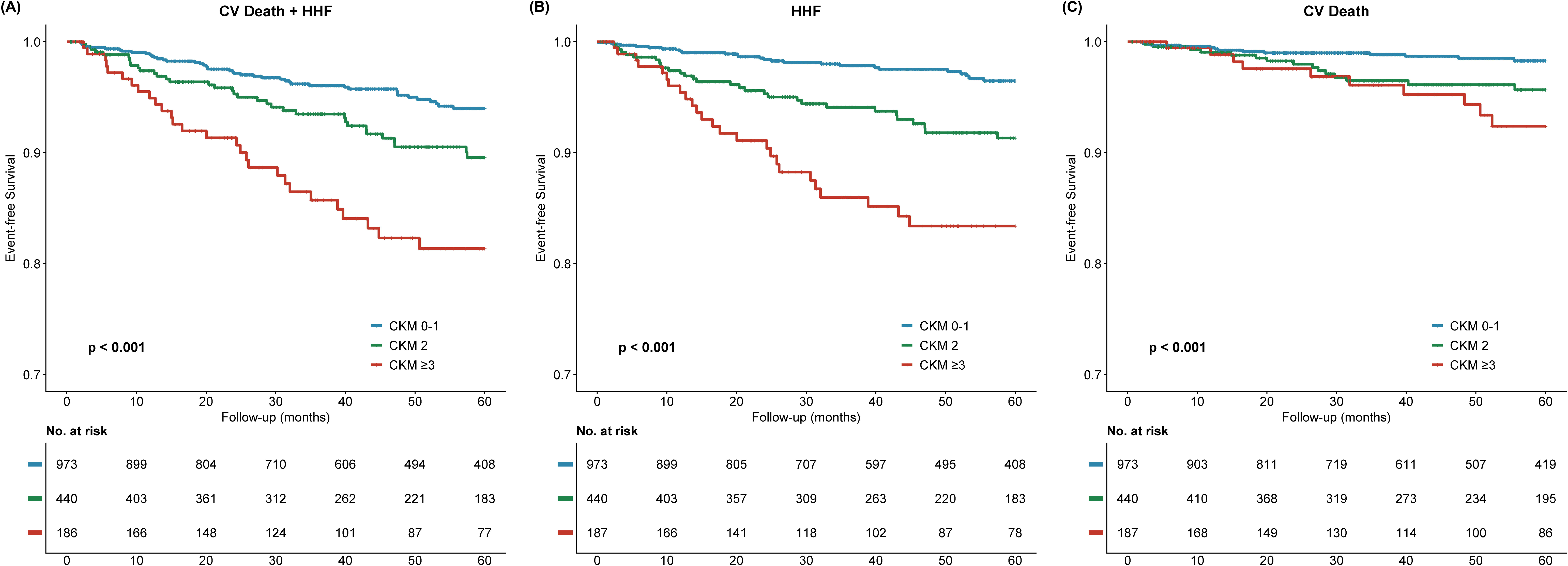
Cardiovascular Outcomes According to CKM Burden Kaplan–Meier curves for cardiovascular outcomes according to CKM burden. (A) composite of cardiovascular death and hospitalization for heart failure (HHF), (B) HHF, and (C) cardiovascular death. Alt text: Three Kaplan–Meier curves comparing cardiovascular outcomes across increasing CKM burden groups. Panels show the composite of cardiovascular death and HHF, HHF, and cardiovascular death, with higher event rates in patients with greater CKM burden.

**Table 3.** Association between CKM burden and cardiovascular outcomes.

| Outcome | Events<br>n (%) | CKM 0–1<br>(ref) | CKM 2<br>HR (95% CI) | P | CKM ≥3<br>HR (95% CI) | P | Per 1-component<br>HR (95% CI) | P |
| --- | --- | --- | --- | --- | --- | --- | --- | --- |
| **Model 1: Adjusted for age and sex** |  |  |  |  |  |  |  |  |
| CV death + HHF | 107 (6.7%) | Ref | 1.84 (1.18–2.87) | 0.007 | 3.20 (1.99–5.15) | <0.001 | 1.54 (1.28–1.85) | <0.001 |
| HHF | 80 (5.0%) | Ref | 2.78 (1.63–4.73) | <0.001 | 5.05 (2.89–8.81) | <0.001 | 1.84 (1.49–2.27) | <0.001 |
| CV death | 38 (2.4%) | Ref | 2.47 (1.17–5.21) | 0.017 | 3.36 (1.46–7.73) | 0.004 | 1.52 (1.11–2.06) | 0.008 |
| **Model 2: Adjusted for age, sex, prior coronary artery disease and stroke** |  |  |  |  |  |  |  |  |
| CV death + HHF | 107 (6.7%) | Ref | 1.82 (1.17–2.84) | 0.008 | 3.09 (1.92–4.98) | <0.001 | 1.53 (1.27–1.84) | <0.001 |
| HHF | 80 (5.0%) | Ref | 2.75 (1.62–4.68) | <0.001 | 4.86 (2.78–8.48) | <0.001 | 1.83 (1.49–2.26) | <0.001 |
| CV death | 38 (2.4%) | Ref | 2.49 (1.18–5.25) | 0.017 | 3.25 (1.41–7.48) | 0.006 | 1.50 (1.10–2.05) | 0.010 |
| **Model 3: Adjusted for age, sex, prior coronary artery disease, stroke and HSI-defined MASLD** |  |  |  |  |  |  |  |  |
| CV death + HHF | 107 (6.7%) | Ref | 1.75 (1.12–2.75) | 0.015 | 2.83 (1.71–4.69) | <0.001 | 1.50 (1.24–1.81) | <0.001 |
| HHF | 80 (5.0%) | Ref | 2.61 (1.52–4.48) | <0.001 | 4.34 (2.41–7.82) | <0.001 | 1.78 (1.43–2.21) | <0.001 |
| CV death | 38 (2.4%) | Ref | 2.52 (1.19–5.36) | 0.016 | 3.36 (1.41–8.05) | 0.006 | 1.48 (1.08–2.05) | 0.016 |

Abbreviations: CKM, cardiovascular-kidney-metabolic; CV, cardiovascular; HHF, hospitalization for heart failure; HR, hazard ratio; CI, confidence interval; HSI, hepatic steatosis index; MASLD, Metabolic dysfunction–associated steatotic liver disease.
Per 1-component HR indicates the risk increment per additional CKM component, modeled as a continuous variable (range 0–4).

In mediation analyses of the association between high CKM burden and the primary outcome, LVGLS showed the largest mediation effect (8.8%; 95% CI, 3.8–18.4%), followed by LVMI (6.3%; 95% CI, 2.8–13.1%) and E/e′ (6.2%; 95% CI, 2.6–13.7%). LASr (3.1%; 95% CI, 0.4–8.6%), and LA stiffness index (3.0%; 95% CI, 1.0–6.7%) also showed statistically significant mediation effects (**Supplementary Table S3**).

### Association of MASLD With CKM Burden and Cardiovascular Outcomes

Given that MASLD represents a hepatic manifestation of metabolic dysfunction closely linked to CKM burden, we additionally evaluated the prognostic relevance of MASLD and whether the association between CKM burden and CV outcomes persisted after accounting for MASLD. HSI-defined MASLD became progressively more prevalent with increasing CKM burden (17.6%, 37.3%, and 51.3% in CKM burden 0–1, 2, and ≥3 groups, respectively; p<0.001). MASLD was associated with a higher risk of the primary composite outcome after adjustment for age and sex (HR, 1.59; 95% CI, 1.03–2.45; p=0.036); however, this association was attenuated after further adjustment for CKM burden and did not reach statistical significance (HR, 1.15; 95% CI, 0.73–1.83; p=0.539). MASLD was not independently associated with longitudinal echocardiographic recovery after adjustment for baseline values, age, sex, and change in SBP. The associations between CKM burden and CV outcomes remained robust after additional adjustment for HSI-defined MASLD (CKM burden ≥3: HR, 2.83; 95% CI, 1.71–4.69; p<0.001, **Table 3**).

## Discussion

In this study, we evaluated the association between CKM burden and CV outcomes in patients with hypertension. The main findings are as follows. First, higher CKM burden was independently associated with an increased risk of adverse CV outcomes. Second, higher CKM burden was associated with the more advanced cardiac remodeling and greater functional impairment at baseline, as well as less favorable recovery of functional echocardiographic parameters, including LVGLS, E/e′ ratio, and the LA stiffness index. Changes in structural parameters, however, were comparable across CKM burden groups. Third, normalization of these functional parameters at follow-up was associated with substantially lower subsequent CV risk. Taken together, these findings suggest that higher CKM burden is associated with impaired recovery of cardiac function after hypertension treatment.

### CKM Burden and Cardiac Functional Recovery

At baseline, increasing CKM burden was accompanied by progressively greater cardiac abnormalities, including higher LVMI, LAVI, E/e′ ratio, and LA stiffness index and impaired LVGLS and LASr. While the association between CKM burden and cardiac abnormalities has been reported,^24^ we expanded the scope to the longitudinal changes in echocardiographic parameters according to the burden of CKM. Following antihypertensive treatment, most echocardiographic parameters improved across CKM burden groups. However, the pattern of recovery differed between structural and functional measures. Although structural measures such as LVMI and LAVI improved similarly across CKM burden groups, LVGLS, E/e′ ratio, and LA stiffness index showed less favorable recovery in patients with high CKM burden. This differential response may reflect distinct determinants of structural and functional cardiac remodeling. Structural changes such as LV hypertrophy are closely linked to hemodynamic load and may therefore improve substantially with BP reduction, irrespective of the accompanying CKM burden.^9,11^ In contrast, functional abnormalities reflected by LVGLS, E/e′ ratio, and LA stiffness index may be influenced not only by elevated BP but also by the broader metabolic and renal abnormalities encompassed by CKM syndrome. Inflammation, oxidative stress, insulin resistance, and vascular dysfunction, which are central pathophysiologic processes in CKM syndrome, may contribute to persistent cardiac dysfunction beyond the effects of elevated BP.^1^ Thus, higher CKM burden may limit the recovery of cardiac function even when structural parameters improve following antihypertensive treatment.

### Prognostic Significance of Echocardiographic Normalization

Echocardiographic normalization was strongly associated with subsequent CV outcomes. Among the three parameters that showed differential recovery across CKM burden groups, LVGLS, E/e′ ratio, and LA stiffness index, patients whose initially abnormal values normalized at follow-up had substantially lower risks of subsequent CV events than those with persistent abnormalities. The proportion of patients achieving LVGLS normalization also progressively decreased with increasing CKM burden. These findings highlight the potential prognostic importance of cardiac functional recovery beyond abnormalities observed at a single time point. Persistent echocardiographic abnormalities despite antihypertensive treatment may represent a marker of residual CV risk, whereas normalization may indicate a more favorable cardiac response. Serial echocardiographic assessment may therefore help characterize cardiac response to treatment and identify patients who remain at increased CV risk despite BP management.

### CKM Burden and Cardiovascular Risk

The concept of CKM syndrome emphasizes the interconnected and multidirectional relationships among metabolic risk factors, kidney dysfunction, and the CV system.^1,2^ The current CKM staging framework provides a clinically useful construct reflecting disease progression, ranging from excess or dysfunctional adiposity to metabolic risk factors and chronic kidney disease and ultimately to subclinical and overt CV disease.^2,8^ However, individuals within the same CKM stage may have substantially different cumulative burdens of obesity, hypertension, diabetes, and kidney dysfunction, reflecting considerable heterogeneity within each stage.

Our findings extend the CKM framework by demonstrating a graded increase in CV risk with increasing cumulative CKM burden among patients with hypertension. Importantly, this association was consistently observed when each CKM component was sequentially excluded from the burden score, indicating that the relationship was not predominantly attributable to any single component. The association also persisted after adjustment for established CV disease and remained robust after additional adjustment for MASLD. These findings suggest that consideration of the number of coexisting CKM abnormalities may provide complementary information regarding CV risk within the broader CKM framework. In addition, serial echocardiographic evaluation may provide further prognostic information by identifying persistent functional abnormalities despite antihypertensive treatment.

### Limitations

Several limitations should be acknowledged. First, this was a retrospective observational study conducted at two tertiary referral centers, and residual confounding and selection bias cannot be excluded. Second, although serial echocardiography allowed assessment of longitudinal cardiac changes following antihypertensive treatment, the retrospective design limited detailed assessment of treatment exposure, including medication classes, treatment intensity, adherence, and their potential differential effects on cardiac remodeling. Third, CKM burden was based on four readily available clinical components with equal weighting and therefore may not fully capture the heterogeneous biological and prognostic contributions of individual CKM domains. Obesity was defined using BMI alone, without measures of body fat distribution or dysfunctional adiposity, and albuminuria was not systematically available, such that the renal component was defined based on eGFR alone. Nevertheless, the association between CKM burden and CV outcomes remained consistent after sequential exclusion of each individual component.

## Conclusion

In patients with hypertension, higher CKM burden was associated with greater cardiac structural and functional abnormalities at baseline, less favorable recovery of functional echocardiographic parameters after antihypertensive treatment, and increased CV risk. Normalization of these functional parameters at follow-up was associated with lower subsequent CV risk, suggesting that less favorable cardiac functional recovery may partly explain the increased CV risk associated with higher CKM burden. These findings support the clinical relevance of the CKM framework for risk stratification and longitudinal management of patients with hypertension.

## Data Availability

The datasets used and analyzed during the current study are available from the corresponding author on reasonable request.

## Acknowledgements

AI-assisted tools were used for manuscript language editing.

## Abbreviations

CKM: cardiovascular-kidney-metabolic
LVGLS: left ventricular global longitudinal strain
CV: cardiovascular
HHF: hospitalization for heart failure
HR: hazard ratio
CI: confidence interval.

