## Supplementary Tables S1-S3, Supplementary Figures S1-S2 for "Impact of Cardiovascular-Kidney-Metabolic Burden on Echocardiographic Remodeling and Cardiovascular Outcomes in Hypertension"

**Supplementary Table S1. Longitudinal changes in echocardiographic parameters according to CKM burden**

| **Parameter** | **CKM Group** | **Baseline** | **Follow-up (Observed)** | **ANCOVA-adjusted EMM** | **p**  **(within group)*** | **p**  **(between group)** **†** |
| --- | --- | --- | --- | --- | --- | --- |
| **LVMI (g/m^2^)** | 0–1 | 105.1 ± 30.8 | 106.5 ± 32.5 | 108.7 ± 0.9 | 0.156 | Ref |
|  | 2 | 116.1 ± 37.7 | 112.9 ± 34.0 | 109.9 ± 1.4 | 0.057 | 0.151 |
|  | ≥3 | 119.4 ± 34.2 | 114.0 ± 35.0 | 109.0 ± 2.1 | 0.034 | 0.349 |
| **LAVI (mL/m^2^)** | 0–1 | 38.6 ± 20.7 | 38.4 ± 19.6 | 39.2 ± 0.4 | 0.592 | Ref |
|  | 2 | 41.4 ± 18.4 | 40.2 ± 17.6 | 39.7 ± 0.6 | 0.066 | 0.234 |
|  | ≥3 | 44.9 ± 36.6 | 43.0 ± 19.2 | 40.1 ± 1.0 | 0.426 | 0.169 |
| **Mitral E/e′ ratio** | 0–1 | 12.2 ± 5.9 | 11.8 ± 5.5 | 12.3 ± 0.2 | 0.022 | Ref |
|  | 2 | 13.3 ± 7.0 | 13.0 ± 6.7 | 13.0 ± 0.2 | 0.198 | <0.001 |
|  | ≥3 | 15.4 ± 7.8 | 15.1 ± 8.8 | 13.6 ± 0.4 | 0.553 | <0.001 |
| **LVGLS (%)** | 0–1 | 16.4 ± 4.7 | 17.1 ± 4.0 | 16.8 ± 0.1 | <0.001 | Ref |
|  | 2 | 14.6 ± 5.1 | 15.9 ± 4.7 | 16.4 ± 0.2 | <0.001 | 0.058 |
|  | ≥3 | 13.8 ± 4.7 | 15.3 ± 3.8 | 16.1 ± 0.3 | <0.001 | 0.036 |
| **LASr (%)** | 0–1 | 29.0 ± 11.7 | 29.4 ± 11.0 | 28.9 ± 0.3 | 0.258 | Ref |
|  | 2 | 27.6 ± 12.2 | 29.4 ± 11.9 | 29.3 ± 0.4 | <0.001 | 0.368 |
|  | ≥3 | 25.3 ± 11.3 | 27.4 ± 10.6 | 29.0 ± 0.6 | 0.006 | 0.833 |
| **LA stiffness index** | 0–1 | 0.59 ± 0.73 | 0.54 ± 0.72 | 0.58 ± 0.02 | 0.004 | Ref |
|  | 2 | 0.73 ± 0.97 | 0.62 ± 0.72 | 0.59 ± 0.03 | 0.001 | 0.374 |
|  | ≥3 | 0.84 ± 0.76 | 0.78 ± 0.94 | 0.67 ± 0.04 | 0.212 | 0.011 |

* P (within group): paired t-test comparing baseline to follow-up values within each CKM burden group.

† P (between group): ANCOVA-adjusted comparison of follow-up values against the CKM burden 0–1 reference group, with baseline value, age, sex, and change in systolic blood pressure as covariates.

Abbreviations: CKM, cardiovascular-kidney-metabolic; EMM, estimated marginal mean; E/e′, mitral E/e′ ratio; LVGLS, left ventricular global longitudinal strain; LASr, left atrial reservoir strain; LA, left atrial; ANCOVA, analysis of covariance; LAVI, left atrial volume index; LVMI, left ventricular mass index.

**Supplementary Table S2. Sensitivity analyses of the association between CKM burden and cardiovascular outcomes after sequential exclusion of individual CKM components**

| **Model** | **CV death+ HHF Adjusted HR (95% CI)** | **p** | **HHF**  **Adjusted HR (95% CI)** | **p** | **CV death Adjusted HR (95% CI)** | **p** |
| --- | --- | --- | --- | --- | --- | --- |
| Full model | 1.55 (1.29–1.85) | <0.001 | 1.84 (1.49–2.27) | <0.001 | 1.52 (1.11–2.07) | 0.008 |
| Excluding obesity | 1.57 (1.28–1.96) | <0.001 | 2.02 (1.60–2.55) | <0.001 | 1.67 (1.18–2.35) | 0.004 |
| Excluding diabetes | 1.49 (1.19–1.86) | <0.001 | 1.78 (1.38–2.31) | <0.001 | 1.33 (0.91–1.95) | 0.146 |
| Excluding CKD | 1.56 (1.24–1.95) | <0.001 | 1.88 (1.45–2.44) | <0.001 | 1.29 (0.88–1.89) | 0.198 |
| Excluding Severe HTN | 1.67 (1.36–2.06) | <0.001 | 1.88 (1.48–2.39) | <0.001 | 1.85 (1.31–2.61) | <0.001 |

Abbreviations: CKM, cardiovascular-kidney-metabolic; CV, cardiovascular; HHF, hospitalization for heart failure; HR, hazard ratio; CI, confidence interval; CKD, chronic kidney disease; HTN, hypertension.

All models adjusted for age and sex.

**Supplementary Table S3. Mediation analysis of the association between high CKM burden and cardiovascular outcomes through echocardiographic parameters**

| **Mediator** | **Proportions Mediated  (95% CI)** | **Indirect effect (ACME) (95% CI)** | **p** |
| --- | --- | --- | --- |
| LVGLS | 8.8% (3.8–18.4%) | 0.012 (0.006–0.020) | <0.001 |
| LVMI | 6.3% (2.8–13.1%) | 0.008 (0.004–0.010) | <0.001 |
| Mitral E/e’ ratio | 6.2% (2.6–13.7%) | 0.008 (0.004–0.014) | <0.001 |
| LASr | 3.1% (0.4–8.6%) | 0.004 (0.001–0.010) | 0.027 |
| LA stiffness index | 3.0% (1.0–6.7%) | 0.004 (0.001–0.008) | 0.002 |

Causal mediation analysis, bootstrap n=5,000. Logistic outcome model. Covariates: age and sex. Exposure: CKM ≥3 vs 0–2. Outcome: CV death or hospitalization for heart failure.

ACME, average causal mediation effect; CI, confidence interval; CKM, cardiovascular-kidney-metabolic; LVGLS, left ventricular global longitudinal strain; LVMI, left ventricular mass index; LASr, left atrial reservoir strain.

**Supplementary Figure S1. Study Flow and Classification According to CKM Burden**

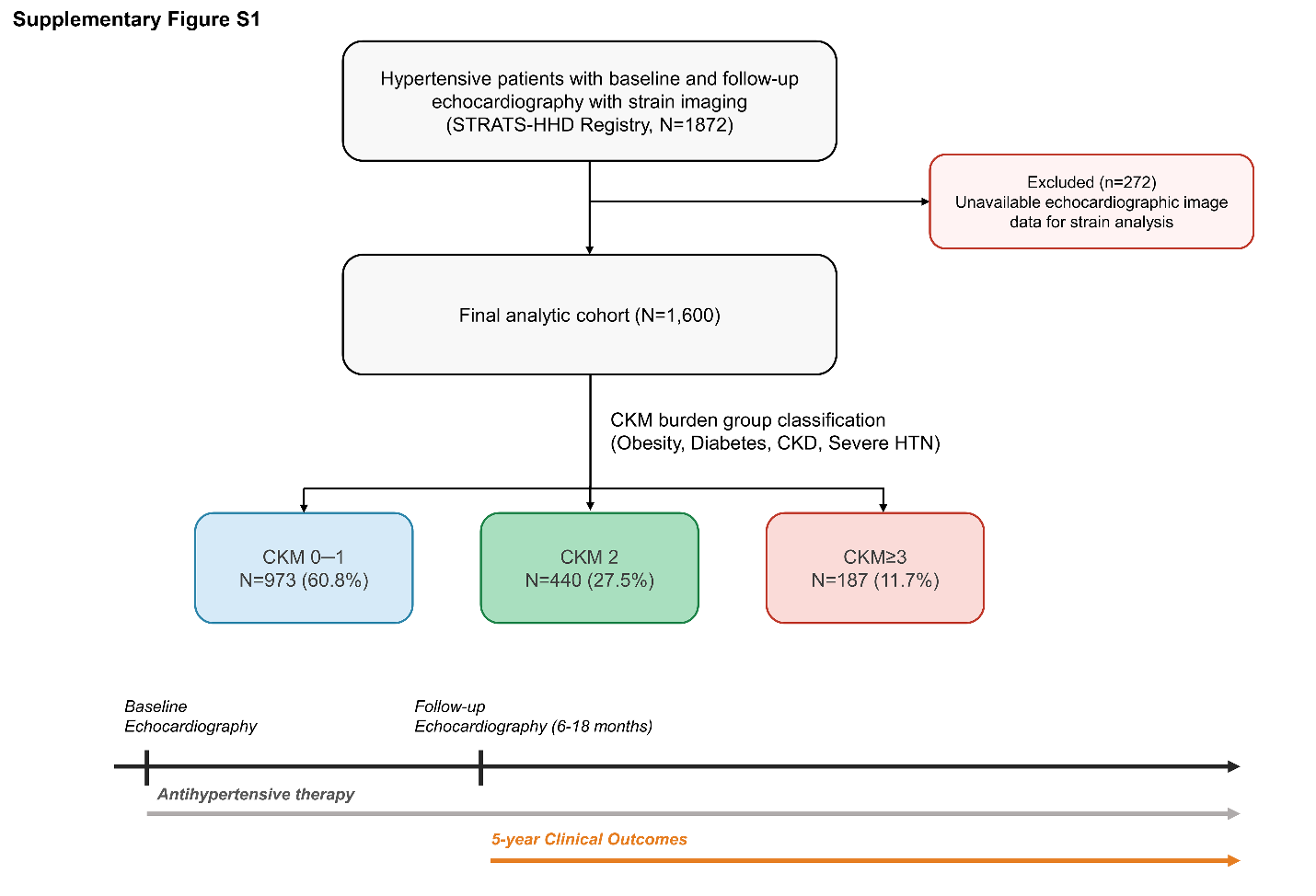

Study flow and classification of patients according to CKM burden. CKM burden was defined as the cumulative number of four components: obesity, diabetes mellitus, chronic kidney disease, and severe hypertension. Patients were categorized into three groups according to the number of CKM components: 0–1, 2, and ≥3.

Alt text: Flowchart showing the selection of 1,600 patients from the STRATS-HHD registry after exclusion of 272 patients, classification into three CKM burden groups (0–1, 2, and ≥3), and the timeline of baseline and follow-up echocardiography and cardiovascular outcome assessment.

**Supplementary Figure S2. Association Between CKM Burden and Cardiovascular Outcomes**

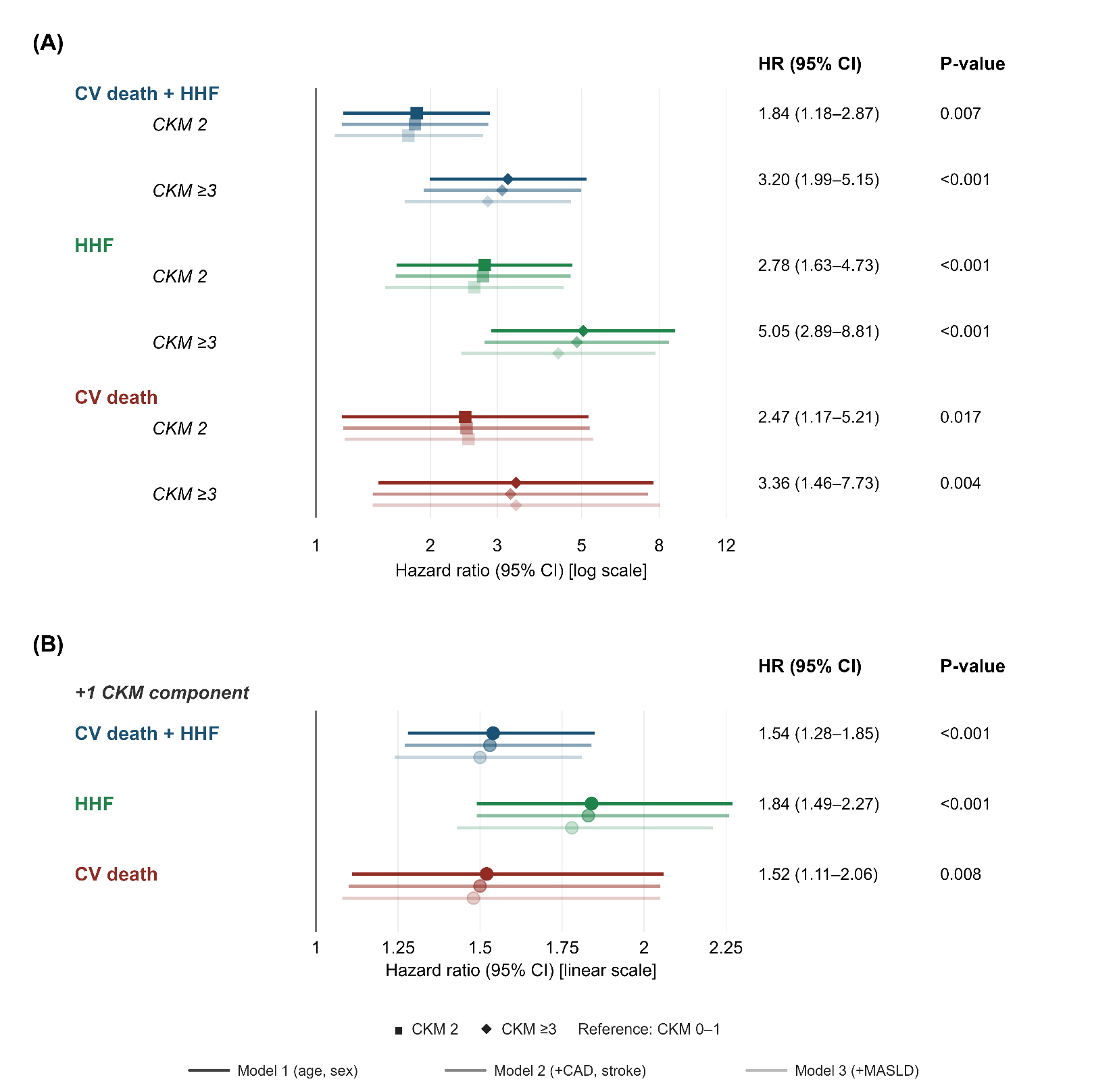

Forest plots showing the associations between CKM burden and cardiovascular outcomes. (A) Hazard ratios (HRs) and 95% confidence intervals (CIs) for CKM burden scores of 2 and ≥3, compared with CKM burden 0–1. (B) HRs and 95% CIs per one-component increase in CKM burden. Three adjustment models were applied: Model 1, adjusted for age and sex; Model 2, additionally adjusted for prior coronary artery disease and stroke; and Model 3, additionally adjusted for hepatic steatosis index-defined metabolic dysfunction-associated steatotic liver disease (MASLD).

Abbreviations: CKM, cardiovascular-kidney-metabolic; CV, cardiovascular; HHF, hospitalization for heart failure; HR, hazard ratio; CI, confidence interval; MASLD, metabolic dysfunction-associated steatotic liver disease.

Alt text: Two-panel forest plot showing higher risks of cardiovascular death, HHF, and their composite with increasing CKM burden. Associations are shown across three adjustment models, both by CKM burden category and per additional CKM component.
